# Effectiveness of a Blended Learning Approach for Strengthening Capacity in Neglected Tropical Disease Control in India and Nigeria

**DOI:** 10.64898/2026.06.29.26356810

**Authors:** Sunday Udo, Joydeepa Darlong, Pravin Kumar, Dinesh Kumar, Mikail Ibrahim, Tabitha Ayuba, Paul Tsaku, Christine Fenenga

## Abstract

**Background:** Persistent gaps in Neglected Tropical Diseases (NTDs) knowledge and skills among frontline health workers in endemic countries remain an important factor limiting progress toward elimination targets. Blended learning approaches offer a potentially scalable solution, but robust evidence from low- and middle-income countries (LMICs) remains scarce. This study evaluated the effectiveness, acceptability, behavioral effects, and cost of a blended learning program for NTD capacity building in India and Nigeria.

**Methods:** A mixed-methods intervention study was conducted in India and Nigeria. A total of 177 purposively selected health workers across three levels (frontline/community workers [Level 1], clinicians and primary care providers [Level 2], and district supervisors/program managers [Level 3]) participated in a blended training program combining interactive online modules with practical onsite skill sessions. Guided by the integrated frameworks of Implementation Science and Kirkpatrick, outcomes were assessed using pre/post knowledge tests, 3–6 month follow-up observations and interviews, focus group discussions, and comprehensive costing. Quantitative data were analyzed using Stata, and qualitative data underwent thematic analysis.

**Results:** The program achieved high acceptability across all cadres and settings. Knowledge scores improved significantly after training (average gains 5–42.5%, p<0.001, Cohen’s d 1.1–1.8) across the 3 training courses. At 3–6 months, workplace observations and supervisor feedback observed improved service delivery. Challenges included internet connectivity, language barriers, and lower online completion rates among Level 1 workers.

**Conclusions:** A contextually adapted blended learning approach is feasible, acceptable, effective, and cost-efficient for strengthening NTD workforce capacity in resource-limited settings. With targeted adaptations (local languages, offline access, and cadre-specific tailoring), this model offers a promising strategy to support the WHO NTD Roadmap 2021–2030 and Zero Leprosy goals.

**Author Summary:** Neglected tropical diseases (NTDs) continue to cause suffering, disability, and stigma for millions of people, especially in countries like India and Nigeria. A major challenge is that many health workers lack up-to-date knowledge and practical skills to diagnose, treat, and support affected people effectively.

In this study, we developed and tested a blended learning approach that combines online courses with hands-on, in-person training. We trained 177 health workers at different levels, from community volunteers to doctors and program managers, in one state in India and two states in Nigeria.

We found that the training was very well received. Participants’ knowledge improved significantly, especially in difficult areas such as managing leprosy reactions and reducing stigma. Three to six months later, health workers were applying what they learned: they were better at examining patients, detecting cases earlier, counselling people with compassion and providing better care. The entire program was delivered on budget, showing it is affordable and practical.

Our findings show that blended learning can be an effective, acceptable, and be delivered at a reasonable cost to strengthen health workers’ skills in resource-limited settings. With some improvements, such as materials in local languages and better offline access, this approach can help many more countries make real progress toward ending NTDs and achieving the global goal of zero leprosy.

## Introduction

Neglected tropical diseases (NTDs), remain a major public health challenge in many low- and middle-income countries (LMICs), such as India and Nigeria. Comprising 21 conditions, NTDs disproportionately affect impoverished populations in tropical and subtropical regions, causing morbidity, disability, stigma, and economic loss [1,2]. These include skin NTDs (e.g., leprosy, Buruli ulcer, yaws, lymphatic filariasis, mycetoma, scabies) and other diseases such as schistosomiasis, soil-transmitted helminthiases, leishmaniasis, and onchocerciasis. Despite progress through mass drug administration, vector control, and improved diagnostics, over 1.5 billion people still require interventions, with ongoing transmission in high-burden LMICs [3,4].

NTDs are strongly associated with long-term disability, stigma, and socioeconomic disadvantage [5–7]. While global efforts have advanced control and elimination, gaps in early detection, diagnosis, and management persist. The WHO NTD Roadmap 2021–2030 emphasizes integrated, people-centered approaches and stronger health systems to achieve prevention, control, elimination, and eradication targets [8,9].

A major constraint is declining health workforce capacity [8–10] Health workers across community, primary, and specialized levels require context-specific skills for diagnosis, case management, disability prevention, stigma reduction, and mental health support [11]. However, training is often limited, irregular, and resource-constrained, with traditional in-person workshops lacking scalability, continuity, and long-term impact.

In response to the dwindling capacity of healthcare workers for integrated NTDs, which poses a significant obstacle to achieving elimination targets the WHO Roadmap 2021-2030, the Lift Leprosy Learning (LLL) project was initiated by member organizations of the International Federation of Anti-Leprosy Organizations (ILEP) in 2021. The LLL mandate encompassed: (i) pooling of global expertise; (ii) advocacy and collaboration among stakeholders; (iii) developing, implementing, updating, and modernizing existing training materials and tools into blended training strategies and materials; and (iv) piloting and evaluating the materials.

Blended learning, combining digital and in-person methods, offers a scalable alternative, improving access, flexibility, and sustained engagement for knowledge retention and skills development [12]. In NTD contexts, it may also support integration of clinical skills and attitudinal change, particularly for stigmatized conditions such as leprosy. Digital tools like the WHO Skin NTDs App can further facilitate integrated learning across conditions [13,14].

Evidence on blended learning for NTDs remains limited, especially across health system levels and high-burden settings. Most studies focus on a single disease or cadre, with little evaluation of integrated or system-level approaches. India and Nigeria illustrate these challenges: India accounts for over half of global new leprosy cases, while Nigeria continues to report transmission despite meeting elimination thresholds. In both countries, workforce constraints hinder early detection, integrated care, and stigma reduction, and evidence for scalable training approaches remains scarce [15].

The CapaBLe (Capacity Building for Leprosy and other NTDs) study emerged from the Lift Leprosy Learning (LLL) aiming to generate evidence on the impact of blended learning interventions across three course topics on NTD-related knowledge and competencies among health workers at three health system levels in India and Nigeria. Leprosy provides a useful model for broader skin NTD training due to its established protocols, focus on disability prevention, and psychosocial care elements transferable to other conditions [16]. By evaluating outcomes across diverse settings and cadres, the study seeks to inform scalable training strategies and monitoring tools for NTD control.

## Methods

### Study design

This was a mixed-methods intervention study conducted between 2024 and 2026 in India and Nigeria, designed to evaluate the effectiveness, acceptability, and cost of blended learning for NTDs health workers. Quantitative assessments (knowledge and skills improvement) and qualitative exploration (perceptions, acceptability, behavioral change) were integrated, enabling triangulation of data.

### Conceptual framework

The study was guided by the Kirkpatrick framework to assess training outcomes across reaction, learning, and behavior levels. For real-world relevance, an implementation science lens was applied to examine embeddedness, acceptability, uptake, fidelity, and sustainability of the blended training approach. The model provides a comprehensive structure to assess not only individual-level outcomes but also the broader contextual factors influencing real-world uptake, delivery, and sustainability of programs. Understanding the barriers and facilitators will allow for co-design strategies with stakeholders to improve uptake [17, 18].

**Figure 1.**
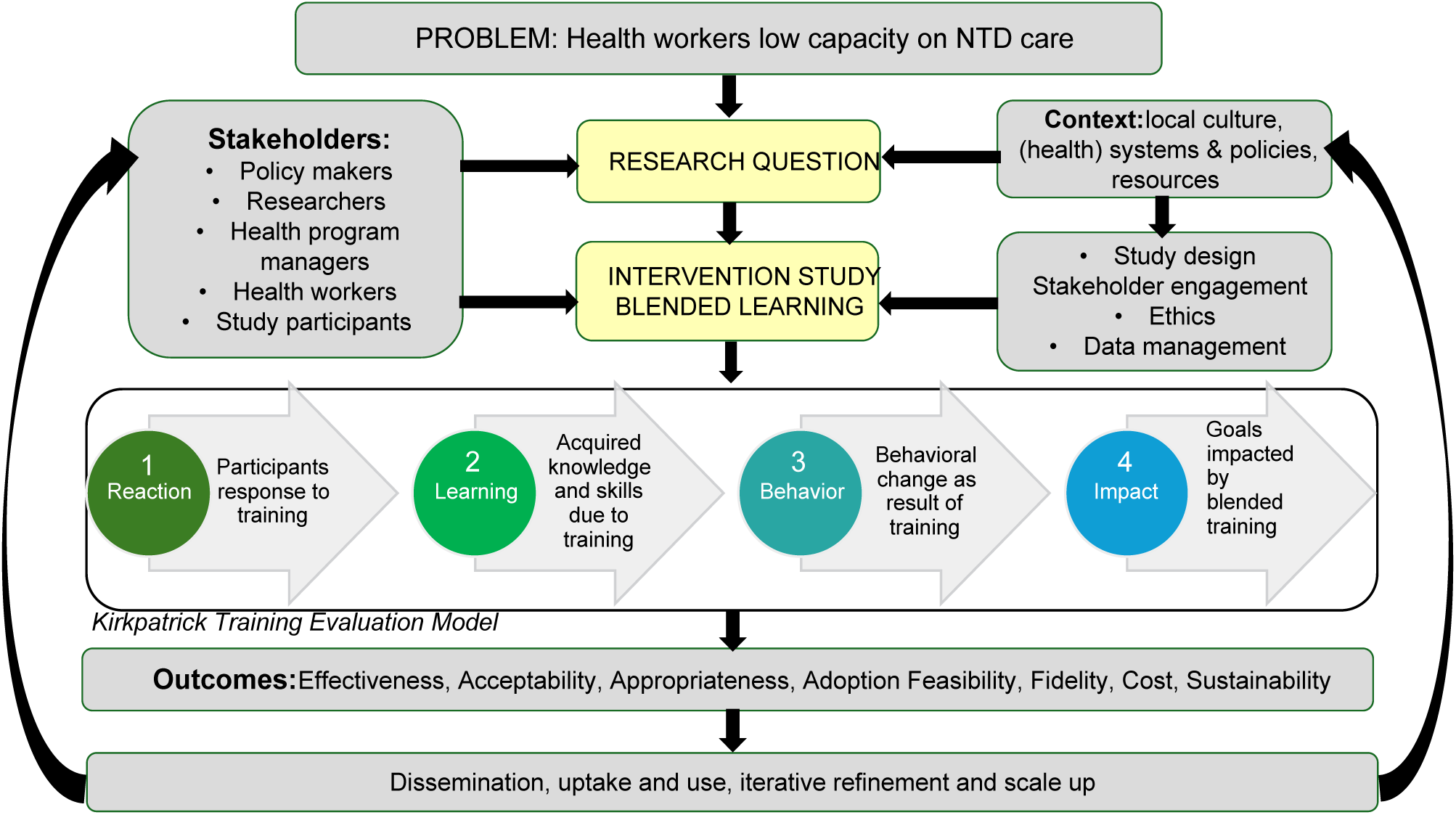
CapaBLe Conceptual Framework – Implementation Science with integrated Kirkpatrick Model.

### Study setting and participants

The study was conducted in high-burden settings for NTDs in two separate areas in Chhattisgarh State, India, and Sokoto and Niger States, Nigeria. These locations were purposefully selected in consultation with local health authorities and training institutions.

A total of 177 health workers participated in the study (India: n = 83; Nigeria: n = 94). Participants included cadres from three levels of the health system: frontline workers and community volunteers, clinicians and medical officers, and district-level programme managers (including district leprosy officers and Local Government Area Tuberculosis and Leprosy Supervisors [LGA-TBLS]).

### Intervention

The intervention was a blended capacity-building programme consisting of three courses: (1) General Leprosy and Skin-NTDs, (2) Mental Wellbeing and Stigma in Neglected Tropical Diseases, and (3) Reactions in Leprosy. These topics were selected based on the findings of a prior capacity-building needs assessment.

Materials included online learning modules hosted on the WHO Academy platform (https://whoacademy.org) for the General Leprosy and Skin-NTDs course, and the infoNTD platform (https://e-learning.infontd.org) for the Mental Wellbeing and Stigma in Neglected Tropical Diseases, and Reactions in Leprosy courses. Supplementary readings and practical demonstration kits were also provided.

Procedures comprised six weeks of structured online learning (combining asynchronous and synchronous sessions), followed by 2–3 days of in-person practical training with expert demonstrations.

The intervention was developed and delivered by subject-matter experts from partner institutions. In-person components were conducted at local training venues in Chhattisgarh State, India, and Sokoto and Niger States, Nigeria. The intervention was tailored to local contexts and aligned with national NTD guidelines. Local training institutions were actively engaged in the planning and delivery.

The programme was implemented between October 2024 and April 2026. Participants were encouraged to apply the learned skills in their workplaces, with structured follow-up support visits conducted at 3 and 6 months post-training. No major modifications were made during implementation.

### Data collection

Qualitative (including focus group discussions – FGD, and semi-structured interviews), pre- and post-training questionnaires and observations, were collected to assess changes and retention in knowledge and competencies before and immediately after and between 3 and 6 months after the intervention. The CapaBLe Conceptual Framework was adopted throughout the study. A breakdown of the data collection process is shown in Appendix 1.

### Data analysis

Quantitative data from pre- and post-test scores were analysed to assess changes in knowledge and skills. Cohen’s d effect size was calculated to compare differences between groups and within groups (pre- versus post-intervention), using Stata software.

Qualitative data were analysed using thematic analysis, guided by the CapaBLe Conceptual framework. This approach aimed to identify key themes related to intervention delivery, uptake, acceptability, and contextual influences.

Interview transcripts were analysed thematically through the following steps: 1) Familiarisation with the data by repeatedly reading the transcripts; 2) Development of a codebook based on the interview guide and existing literature; 3) Systematic coding of the transcripts using NVivo software; 4) Identification of additional emergent themes; 5) Summarisation and refinement of each theme; and 6) Interpretation of the findings in relation to the study objectives.

### Validation and Trustworthiness

To enhance the rigour and trustworthiness of the findings, multiple validation strategies were employed across the study. The intervention modules, pre-post-test questions, observation checklists, and interview guides were collaboratively developed and contextualised by a team of seasoned professionals with expertise in both Indian and Nigerian settings. These tools underwent pilot testing and iterative refinement to strengthen their content validity, cultural appropriateness, and reliability.

During data collection, co-researchers systematically reviewed all quantitative and qualitative data for completeness and accuracy. For the qualitative component, the entire dataset was coded systematically and consistently using NVivo, with dual independent coding applied to a sample of transcripts to enhance dependability and confirmability. Thematic summaries were validated through repeated returns to the original transcripts. Interpretations were cross-checked against the CapaBLe Conceptual Framework, existing literature, and through “expert checking” by site co-researchers who acted as cultural brokers. Member-checking with selected participants was also conducted where feasible. An audit trail of analytical decisions, reflexive memos, and team discussions was maintained throughout the process. Direct participant quotes are presented in the results to allow readers to judge the credibility and authenticity of the interpretations.

### Dissemination for uptake and use

Findings from the study are actively disseminated to promote uptake and inform policy and practice. This includes targeted presentations of key results to local and global stakeholders, such as study participants, Ministries of Health in India and Nigeria, implementing partners, and relevant development agencies.

A concise, accessible policy brief has been developed and is being shared with decision-makers and other stakeholders. In addition, the results are disseminated through peer-reviewed open-access publications, stakeholder workshops, and targeted knowledge translation activities. These efforts aim to facilitate the translation of evidence into actionable improvements in capacity-building interventions for health professionals.

### Ethical considerations

Ethical approvals were obtained from the appropriate ethics committees in both India and Nigeria. In India, approval was granted by the LEPRA Society Institutional Ethics Committee in Hyderabad (reference number: 04/LEPRA IEC/2024-2025). In Nigeria, approval was granted by the National Health Research Ethics Committee (NHREC) in Abuja (reference number: NHREC/01/01/2007-18/11/2024). These approvals adhered to the Declaration of Helsinki. Participants for the interventions were recruited through collaborating health institutions and training programs, and written informed consent was obtained from all participants.

## Results

### Study Participants

A total of 177 participants were recruited across the two study sites, comprising 83 participants from India and 94 participants from Nigeria. Participants were categorized into three training levels based on their roles within the health system: Level 1 (community-level workers/volunteers), Level 2 (community and primary healthcare providers), and Level 3 (medical officers, specialists, and health supervisors). The characteristics of participants by country and training level are summarized in Table 1.

**Table 1.** Characteristics of study participants by country and training level.

| Level | India (n=83) | Nigeria (n=94) |
| --- | --- | --- |
| **Level 1** | **33 mitanin (ASHA) workers** involved in community health promotion and case detection. All completed in-person training; online modules delivered in Hindi but no pre/post-test data due to low literacy. Mitanins are key for community engagement and stigma reduction. | **38 community volunteers** (21 Sokoto, 17 Niger). All completed in-person training; 35 enrolled in online courses. Volunteers support first aid, MDA, and referral of people affected by NTDs. |
| **Level 2** | **27 community/rural health providers**<br>Community Health Officers (n = 12, 44.4%), Rural Health Officers (n = 11, 40.7%), and Rural Medical Assistants (n = 4, 14.8%). All attended in-person training. Online completion ranged from **63–74%**, with some participants blocked after three failed attempts. Main challenge: online assessment restrictions. | **37 healthcare workers**<br>The participants consisted of medical doctors (n = 7, 18.9%), nurses (n = 19, 51.4%), and community health extension workers (n = 11, 29.7%). All completed in-person training; 30, 25, and 20 enrolled in the three online modules. Reduced online participation was mainly due to poor internet access and lack of suitable devices. |
| **Level 3** | **23 medical officers/specialists**, predominantly Medical Officers. In-person participation was high; assessment completion was **65–70%**. Online completion was limited by the three-attempt restriction. | **19 health supervisors** from state ministries of health. All completed in-person training; **13 participated online**. Lower online participation was attributed to poor internet connectivity. |
| **Overall findings** | High acceptance of in-person training across all levels. Online learning was constrained by assessment restrictions and participant completion. | High acceptance of in-person training across all levels. Online participation was mainly limited by connectivity and device availability. |

### Learning Evaluation (Kirkpatrick’s Four-Level Training Evaluation Framework)

The evaluation findings of the training program are presented below using Kirkpatrick’s Four-Level Training Evaluation within our CapaBLe Framework. This widely recognized model provides a structured, progressive approach to measuring training effectiveness at four distinct levels:

- Level 1: Reaction – Participants’ immediate feedback, satisfaction, and perceived value of the training experience.
- Level 2: Learning – The extent to which participants acquired new knowledge, skills, and attitudes during the program.
- Level 3: Behavior – The degree to which participants applied their learned competencies in the workplace.
- Level 4: Results – The organizational impact, business outcomes, and overall return on investment resulting from the training.

The following sections detail the results for each level, supported by quantitative and qualitative data, key observations, and relevant performance metrics.

#### Level 1: Reaction (Acceptability & Satisfaction)

At this level, participants were assessed on their perceptions of the training, including its relevance, content quality, delivery, and whether expectations were met. This result was elicited through a focus group discussion up to two weeks into the training and semi-structured interviews at 3 months. Comparative insights from Indian Level 2 (frontline health workers) and Level 3 (primarily Medical Officers) participants, drawn from detailed semi-structured interviews, provide valuable cross-context understanding of the blended leprosy training program.

##### Overall Reactions and Expectation Fulfillment

In Nigeria, reactions overall were positive across all groups, with participants perceiving the blended approach as accessible, relevant, and exceeding expectations in building foundational knowledge on leprosy, stigma, and mental health.

Most participants reported that the training met or exceeded expectations. For instance, in Level 1 Group 1, Respondent 3 noted,

> *“My expectation was to be able to tell clearly and identify a leprosy case and what type of case it is whether multibacillary or paucibacillary and my expectations were met.”*

In Level 2 Group 1, Respondent 7 expressed gratitude, stating the training met 100% of expectations and provided a certificate for professional value. Level 3 participants, like Respondent 5, said:

> *“You made the training participatory and you have met more than my expectations especially the stigma and mental wellbeing course because before now, I did not know much about it.”*

These views were strongly echoed in India. In the Indian Level 2 cohort, all the participants expressed overall satisfaction. Participants repeatedly described the training as “Very good… modules include all important things.” One L2 participant (L2011) called it *“One of the best trainings for leprosy, with excellent structure and content on reaction and stigma.”* In the Indian Level 3 cohort, most of the participants rated the training as good or very good. Participant L3005 shared:

> *“After the training I felt in myself that I can recognize all the patients very well… I have learnt to identify that very well.”*

Participant L3001 added:

> *“This training was very good. The best thing is that you people did it offline and prepared the basics. On top of that you provided it online also in a systematic and orderly manner, so that we could study anytime…”*

Perceptions in both countries emphasized the training’s role in filling knowledge gaps, with newcomers appreciating introductory content and experienced participants valuing refreshers.

##### Content Relevance and Quality

In Nigeria the content relevance and quality of the materials were perceived as “well detailed,” “self-explanatory,” and “easy to understand.” Level 1 Group 1’s Respondent 2 praised the arrangement for teaching stigma counseling and contact tracing. In Level 2 Group 2, Respondent 2 called it “well explanatory.” Level 3’s Respondent 1 highlighted global relevance and high-quality materials.

A representative Nigerian quote (Level 2 Group 2, Respondent 2) states:

> *“I think it is well detailed because I understand the treatment type, duration, the type of leprosy… The content of the training is well arranged and in order and properly taught. Now I know more about stigma and how to counsel someone with leprosy. I also know more about contact tracing.”*

Indian participants similarly praised content depth and practical value. In Level 2, participants highlighted improved knowledge on reactions and mental health:

> *“We have been studying about reaction due to Leprosy for many days and the stigma in it is even better for our mental health…my knowledge has increased”* (L2_001).

Level 3 Indian participants valued clinical focus and patient demonstrations. Participant L3004 noted:

> *“The one that in which patients were shown, that was the best part… this has happened in this training, which I felt very good.”*

Participant L3008 appreciated learning to differentiate reactions:

> *“I liked the drug reaction i.e., type one and type two Lepra reaction… I started to differentiate from there only.”*

Both cohorts noted the modules helped shift focus from pure physical symptoms to holistic patient care, including mental wellbeing and stigma reduction.

##### Perceptions on Delivery and Mode of Blended Learning

The blended learning approach was effective for its flexibility (online self-pacing) and interaction (in-person). Positive experiences in Nigeria included interactive sessions feeling “equal” and “participatory” (Level 3 Respondent 3).

Indian participants also strongly endorsed the blended model. Most level 2 participants particularly appreciated in-person sessions for peer support and practical skill development:

> *“The things that we do not understand, then we ask our colleagues…there I understand it.”*

Online components were valued for flexibility:

> *“In online sir, we can take our time and properly learn… there are videos as well as written work in it.”* (L2_0011).

The Indian Level 3 participants similarly highlighted the value of combining offline practical exposure with online self-study materials.

##### Challenges and Suggestions for Improvement

Common challenges in Nigeria included network issues (Level 1 Group 2 Respondent 7), short duration (suggestions for 3-5 days), and access barriers. Suggestions included providing manuals/booklets, extending days, translating to Hausa, and adding accommodations.

> *“Yes, there is a challenge with the network and internet. I believe it is a general problem and there is not much we can do concerning that. Asides that, everything else seems perfect.”* (Respondent 1, Level 3, Nigeria)

Similar challenges appeared in India. Language barriers were prominent, with some level 2 participants noting difficulties with English content and recommending Hindi versions:

> *“We don’t know English sir, it should be in Hindi.”* (L2_0012).

Time management, workload, and the need for longer practical sessions were frequently mentioned. Indian participants suggested offline modules, bilingual content, more videos/cartoons, and regular refreshers every 2–6 months. Gender balance was also raised in Nigeria (Level 1 Group 1 Respondents 2 and 3):

> *“I notice that in this selection, there are more males than females and I believe it should be balanced because… males cannot examine the female patients. So, I believe that we should have more females…”*

##### Overall Satisfaction

Overall satisfaction was very positive in both countries. High enthusiasm was evident, with participants expressing gratitude to organizers and a strong sense of empowerment. In Nigeria, the training was well received. In India, approximately 83% of Level 2 and 87.5% of Level 3 participants rated the training between excellent and good. Certificates and structured content served as important motivators in both contexts.

#### Level 2 Learning (Knowledge and Skills Acquisition)

The level 2 of the Kirkpatrick’s framework of learning evaluation focus on measuring what participants actually learned from the training program. It evaluates increase in knowledge, skills, and attitude resulting from the training. In this study, the pre- and post-test was conducted for the participants. The result is presented in table 2 below. Significant improvement was observed in all participant groups across the three courses (General Leprosy and Skin-NTDs, Stigma & Mental Wellbeing, and Reaction in Leprosy). Average score improvements ranged from 5% to 42.5%, where all were statistically significant (p < 0.001) with large effect sizes (Cohen’s d = 1.1 – 1.8), indicating strong training effectiveness.

**Table 2.** Assessment scores (averages) in pre- and post-test.

| Course | Level | Group | N | Pre-test (%) | Post-test (%) | Improvement (%) | p-value (Cohen's d) | Notes |
| --- | --- | --- | --- | --- | --- | --- | --- | --- |
| General leprosy and skin-NTDs | 1 | Nigeria A | 21 | 44.00 | 68.00 | 24.00 | <0.001 (1.2)* | In-person |
|  | 1 | Nigeria B | 17 | 38.00 | 50.00 | 12.00 |  |  |
|  | 2 | Nigeria A | 19 | 54.00 | 73.00 | 19.00 |  |  |
|  | 2 | Nigeria B | 18 | 48.00 | 53.00 | 5.00 |  |  |
|  | 2 | India | 27 | 63.40 | 86.20 | 22.80 |  |  |
|  | 3 | Nigeria A | 12 | 59.00 | 86.00 | 27.00 |  |  |
|  | 3 | Nigeria B | 7 | 39.00 | 67.00 | 28.00 |  |  |
|  | 3 | India | 23 | 73.40 | 86.20 | 12.80 |  |  |
| Stigma & Mental Wellbeing | 1 | Nigeria A | 17 | 73.06 | 90.57 | 17.51 | <0.001 (1.1)** | Online |
|  | 1 | Nigeria B | 18 | 62.44 | 80.94 | 18.50 |  |  |
|  | 1 | Nigeria A | 21 | 75.33 | 91.47 | 16.14 |  | In-person |
|  | 1 | Nigeria B | 17 | 70.50 | 94.11 | 23.61 |  |  |
|  | 2 | Nigeria A | 16 | 74.50 | 86.67 | 12.17 |  | Online |
|  | 2 | Nigeria B | 14 | 75.71 | 86.86 | 11.15 |  |  |
|  | 2 | India | 27 | 50.20 | 88.70 | 38.50 |  |  |
|  | 2 | Nigeria A | 19 | 75.08 | 93.28 | 18.20 |  | In-person |
|  | 2 | Nigeria B | 18 | 80.54 | 97.77 | 17.23 |  |  |
|  | 3 | Nigeria A | 6 | 69.50 | 90.00 | 20.50 |  | Online |
|  | 3 | Nigeria B | 7 | 60.86 | 76.29 | 15.43 |  |  |
|  | 3 | India | 23 | 63.40 | 94.30 | 30.90 |  |  |
|  | 3 | Nigeria A | 12 | 72.34 | 92.05 | 19.71 |  | In-person |
|  | 3 | Nigeria B | 7 | 72.43 | 91.55 | 19.12 |  |  |
| Reaction in leprosy | 2 | Nigeria A | 6 | 29.83 | 72.33 | 42.50 | <0.0001 (1.8)*** | Online |
|  | 2 | Nigeria B | 14 | 56.86 | 84.85 | 27.99 |  |  |
|  | 2 | India | 27 | 50.20 | 88.70 | 38.5 |  | In-person |
|  | 3 | India | 23 | 63.40 | 94.30 | 30.9 |  |  |

\*Participants significantly improved after the training. The effect size shows the training had a strong impact on knowledge or skills.
\*\*Despite a high baseline (71.4%), participants still improved significantly. The training was effective even for those starting with higher knowledge.
\*\*\*This course had the lowest pre-test score but the highest improvement. The training was exceptionally effective, possibly because it covered new or unfamiliar content.

#### Kirkpatrick Level 3: Behavior (On-the-Job Application)

This level measures the extent to which participants apply what they learned during training when they return to their actual work environment. It evaluates behavior change — whether the knowledge, skills, or attitudes acquired in the training are being transferred and used on the job. This assessment was conducted through follow-up visits by the research team 3–6 months after the training intervention. The evaluation included observation of work performance, supervisor/manager feedback, self-assessment by participants, and interviews with trainees and their supervisors.

##### Observation of Work Performance

In both countries, the healthcare workers demonstrated clear improvements in clinical practice. They adopted systematic examination techniques, including proper monofilament testing for sensory loss, nerve palpation, and thorough head-to-toe screening. Participants showed increased confidence in early case detection, differentiating leprosy from other skin conditions, identifying reactions, and making timely referrals. Community-level workers applied active case finding through door-to-door surveys and opportunistic screening.

##### Supervisor/Manager Feedback

Supervisors reported positive behavioral changes among trained staff. They observed better case identification, improved coordination, more confident handling of cases, and proactive knowledge sharing.

The supervisors made remarks such as:

> *“If they suspect a patient in the field, they contact us without hesitation.” –* L3-1 Supervisor, India
>
> *“They make village announcements, conduct surveys, identify patients, and call them for treatment.”* – L3-4 Supervisor, India
>
> *“After the training, they have learned a lot… They are using it properly and linking leprosy work with national programs.”* – L3-3 Supervisor, Nigeria

Supervisors also noted improved patient counseling and reduced treatment discontinuation due to better reaction management.

##### Self-Assessment by Participants

Participants across all three levels from both countries reported actively applying the training in their daily work. They highlighted improved diagnostic skills, reaction management, stigma reduction through counseling, and community awareness activities.

> *“I introduced a WhatsApp group where the local government supervisors send us pictures relevant to the diagnosis of skin diseases, usually the most experienced people amongst us make decisions and sometimes, the patients are sent for examination.”* – Level 2 participant, Nigeria.
>
> *“Previously we would consider any visible patch… as leprosy… now we can identify accurately… I created a patient support group… counselled families… support them psychologically.”* – L2 Participant, India

Many participants also mentioned using the monofilament test instead of a pen and providing better counseling on reactions and treatment adherence.

#### Kirkpatrick Level 4: Results (Broader Impacts)

The 4th level of the Kirkpatrick framework evaluates the overall impact of the training on organizational and programmatic outcomes. It seeks to determine whether the training produced measurable value and contributed to the broader success of the health system or NTD program. In essence, it answers the question: “Did the training deliver tangible results for the organization?”

Due to the relatively short duration of the CapaBLe study, a full assessment of long-term organizational impact was not possible. However, anecdotal evidence and stakeholder feedback indicated several positive outcomes, including:

- Improved integration of NTDs and mental wellbeing services, with health facilities shifting from a siloed disease-specific approach to more holistic patient care.
- Reduction in misdiagnosis and better clinical management leading to improved healing outcomes.
- Increased community sensitization activities and greater professional confidence among healthcare workers.

### Costing

The total cost of implementing the blended learning intervention for healthcare workers managing Neglected Tropical Diseases (NTDs) in India and Nigeria was €103,711.63. This was remarkably close to the planned budget of €107,690, representing a variance of only €3,978.37 (3.7% under budget). See table 3 for the breakdown of the cost. Note that this cost overview excludes development cost of the online and in-person training.

**Table 3.** The cost of implementing a blended learning intervention for NTD workforce in the CapaBLe study in India and Nigeria.

| COMPONENTS | INCLUDES | Nigeria | India | Nigeria | India |
| --- | --- | --- | --- | --- | --- |
|  |  | BUDGET (€) |  | ACTUAL (€) |  |
| **Development costs** | · Planning project | 1,000.00 | 600.00 | 2,500.00 | 661.00 |
|  | · Obtaining ethical approval | 200.00 | 1,500.00 | 93.63 | 1,500.00 |
|  | · Communication | 1,140.00 | 1,140.00 | 1,140.00 | 1,140.00 |
| **Implementation costs** | · Implementing courses | 10,770.00 | 3,000.00 | 10,770.00 | 1,864.00 |
|  | · Follow up field visits | 3,000.00 | 4,000.00 | 3,000.00 | 4,000.00 |
|  | · Analyses of results | 2,100.00 | 4,000.00 | 2,000.00 | 3,531.00 |
|  | · Dissemination of findings | 7,200.00 | 1,000.00 | 7,000.00 | - |
| **Control costs** | · Coordination | 37,176.00 | 20,376.00 | 36,576.00 | 20,376.00 |
|  | · Monitoring and evaluation | 1,500.00 | 1,000.00 | 1,000.00 | 629.00 |
|  | Other overhead costs not foreseen in the above | 3,872.00 | 3,116.00 | 3,872.00 | 2,059.00 |
| **TOTAL** | **Overall costs** | **67,958.00** | **39,732.00** | **67,951.63** | **35,760.00** |

## Discussion

This study provides compelling evidence that a contextually adapted blended learning intervention can effectively strengthen healthcare worker capacity for Neglected Tropical Diseases (NTDs), in two high-burden LMIC settings. Guided by the CapaBLe framework, the intervention achieved consistently positive outcomes across all four Kirkpatrick evaluation levels: high participant satisfaction and acceptability (Level 1), substantial improvements in knowledge and skills with large effect sizes (Level 2), clear transfer of learning into workplace practice (Level 3), and promising early organizational benefits (Level 4), all delivered according to the budgeted costs.

Participants in both India and Nigeria highly valued the blended approach, appreciating the flexibility of self-paced online modules combined with practical in-person skill sessions. The training was perceived as relevant, comprehensive, and transformative, particularly in addressing previously neglected areas such as leprosy reactions, stigma, and mental wellbeing. These findings align with existing literature on blended learning in several contexts, which consistently reports higher learner satisfaction and better knowledge retention compared to traditional single-mode training. The significant pre- to post-test improvements observed across all courses, especially the large gains in the Reactions in Leprosy module, further reinforce evidence that blended formats are well-suited for complex clinical and attitudinal content in NTD programs [19–22].

Notable differences emerged between the two countries. In India, higher literacy levels and relatively better digital access among mid- and senior-level workers facilitated smoother online engagement, although language barriers (English vs Hindi) remained a challenge for frontline cadres. In Nigeria, internet connectivity, device access, and workload issues posed greater obstacles, particularly for Level 1 community volunteers, resulting in lower online completion rates. Despite these contextual variations, strong in-person components, trainer support via WhatsApp, and local adaptations enabled positive outcomes in both settings. This highlights the importance of flexible, hybrid designs that can be tailored to local infrastructure and workforce realities.

At the behavioral level (Kirkpatrick Level 3), both self-reports and supervisor feedback confirmed meaningful on-the-job application, including improved diagnostic practices (e.g., monofilament testing), better reaction management, enhanced patient counselling, and increased community sensitization. These changes contributed to early organizational benefits (Level 4), such as reduced misdiagnosis, more holistic patient care, and stronger integration of mental wellbeing into NTD services. Although there is a paucity of literature on the adoption of the framework in a similar context, the existing reports corroborate these findings [23,24]. Studies such as Laronze & Nkaoua (2024) have identified in their reviews some publications that showed blended learning to have better outcomes than conventional learning. Their findings further revealed a tendency for superior knowledge acquisition in blended learning compared to a single method, higher levels of cognitive engagement, spatial presence and anxiety in in-person than synchronous distance learning, and higher levels of enjoyment for synchronous distance learning [25].

The total implementation cost of €103,711.63, delivered almost exactly on budget (3.7% variance), demonstrates strong financial efficiency and underscores the cost-effectiveness of blended learning as a scalable and sustainable alternative to repeated traditional in-person workshops. These findings are particularly relevant in the context of declining global funding for NTDs. In line with this, researchers have consistently advocated for more cost-effective and innovative approaches to NTD workforce capacity building and program implementation [26,27].

Several challenges and limitations should be acknowledged. Language barriers, text-heavy content, AI voice accents, strict assessment rules (three attempts), and inconsistent internet connectivity hindered online completion, especially among lower-cadre workers. Workload pressures and short in-person duration were also frequently cited. The relatively short follow-up period (3–6 months) limited our ability to fully evaluate long-term organizational and patient-level outcomes (Kirkpatrick Level 4). Furthermore, the absence of a control group restricts definitive causal attribution of observed changes solely to the intervention. These challenges are in tandem with other reports from different contexts. For instance, in Rwanda, for instance, 71.1% of students identified high internet costs and 63.2% cited poor connectivity as major obstacles [28], while some students face computer proficiency challenges, limiting their ability to engage effectively with digital learning materials [29,30], and uneven access to technologies and financial constraints in purchasing data or devices create significant equity gaps [28].

Despite these limitations, the study offers valuable opportunities for advancing NTD workforce development. The strong emphasis on stigma reduction and mental wellbeing integration addresses a critical gap in current programming. Future programs should prioritize local language versions, offline functionality, more visual and interactive content, cadre-specific tailoring, and longer practical sessions. Regular refresher training and structured mentoring mechanisms will be essential to sustain behaviour change. Additionally, the authors agree with the recommendation by Sareen & Mandal (2024), which suggested the adoption of tailored, region-specific policy interventions to mitigate blended learning challenges [31].

## Conclusion

The study demonstrates that a contextually adapted blended learning approach is a feasible, acceptable, and effective strategy for strengthening healthcare worker capacity in neglected tropical diseases, particularly leprosy, across diverse LMIC settings. Guided by the Capable Conceptual Framework, with integrated Implementation Science and Kirkpatrick frameworks, the intervention achieved high participant satisfaction, substantial improvements in knowledge and skills, transfer of learning into clinical practice, and promising early organizational benefits. The combination of self-paced online modules and targeted in-person practical sessions proved especially valuable in addressing both technical competencies and cross-cutting issues such as stigma reduction and mental wellbeing, despite challenges related to digital access, language, and literacy. The Capable Conceptual framework has shown to be a very suitable framework for further research on the effects of training.

These findings underscore the potential of blended learning to support scalable and sustainable capacity building aligned with the WHO NTD Roadmap 2021–2030 and “Towards Zero Leprosy” targets. With further adaptations, including local language versions, enhanced offline functionality, more visual content, and structured follow-up mechanisms, the study model offers a promising pathway for strengthening integrated NTD responses in high-burden countries. Wider adoption and long-term evaluation are recommended to maximize impact on workforce development and elimination goals.

## Data Availability

All relevant data are within the manuscript and its Supporting Information files.

## Acknowledgements

We gratefully acknowledge the resilient people affected by NTDs, who remain key stakeholders in achieving global targets for disease elimination. We also extend our sincere thanks to the members of the LLL steering committee who developed some of the training courses, as well as the CapaBLe Research Advisory Committee: Dr. Benedict Quao, Dr. Helen Roberts, Prof. Zubairu Iliyasu, Dr. David Prakash, and Dr. Obioma Akaniro, for their valuable guidance and contributions. Finally, we deeply appreciate the dedication of health workers across both countries, especially the participants of the CapaBLe study in India and Nigeria, whose commitment continues to drive progress towards ending the neglect of NTDs.

## CRediT Author contributions

**Conceptualization**: Sunday Udo, Joydeepa Darlong, Pravin Kumar, Paul Tsaku, Christine Fenenga

**Data curation**: Sunday Udo, Joydeepa Darlong, Pravin Kumar, Dinesh Kumar, Mikail Ibrahim, Tabitha Ayuba, Paul Tsaku, Christine Fenenga

**Formal analysis**: Sunday Udo, Joydeepa Darlong, Pravin Kumar, Dinesh Kumar, Mikail Ibrahim, Tabitha Ayuba, Paul Tsaku, Christine Fenenga

**Funding acquisition**: Sunday Udo, Joydeepa Darlong, Pravin Kumar, Christine Fenenga

**Investigation**: Joydeepa Darlong, Pravin Kumar, Dinesh Kumar, Mikail Ibrahim, Tabitha Ayuba, Paul Tsaku

**Methodology**: Sunday Udo, Joydeepa Darlong, Pravin Kumar, Paul Tsaku, Christine Fenenga

**Project administration**: Sunday Udo, Joydeepa Darlong, Pravin Kumar, Mikail Ibrahim, Paul Tsaku, Christine Fenenga

**Resources**: Sunday Udo, Joydeepa Darlong, Paul Tsaku, Christine Fenenga

**Software**: Sunday Udo, Joydeepa Darlong, Paul Tsaku

**Validation**: Sunday Udo, Joydeepa Darlong, Pravin Kumar, Dinesh Kumar, Mikail Ibrahim, Tabitha Ayuba, Paul Tsaku, Christine Fenenga

**Writing – original draft**: Joydeepa Darlong, Paul Tsaku, Christine Fenenga

## Appendix 1. Data collection on the effects of intervention

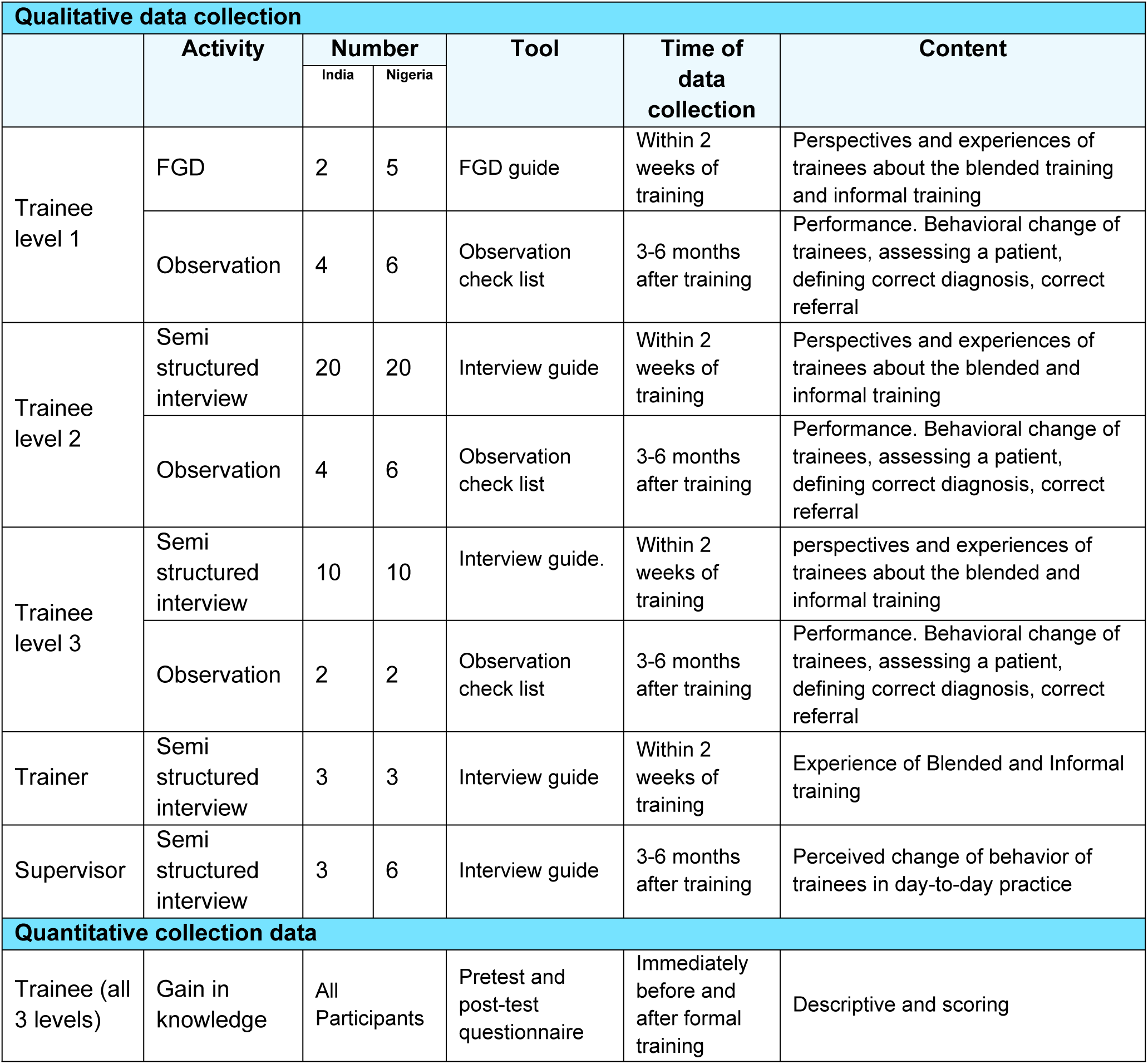
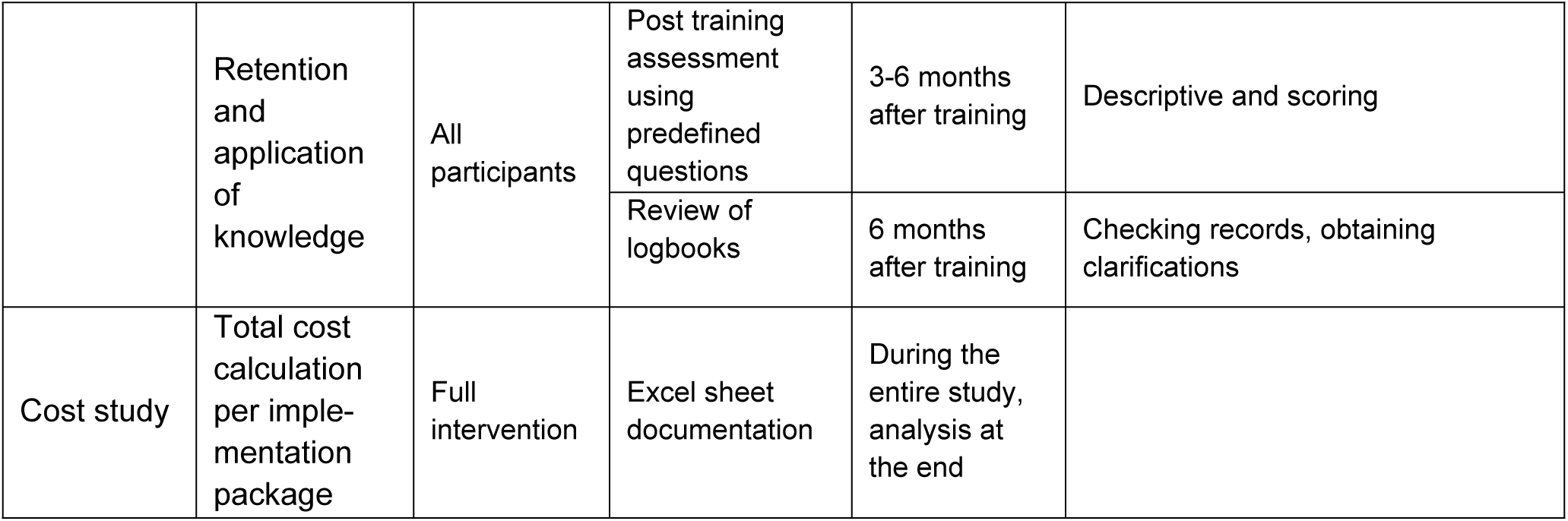

## References

1. Neglected tropical diseases -- GLOBAL [Internet]. [cited 2026 May 5]. Available from: https://www.who.int/health-topics/neglected-tropical-diseases

2. Jesudason T. Global progress report on neglected tropical diseases. Lancet Infect Dis. 2024 Jul 1;24(7):e420. doi:10.1016/S1473-3099(24)00365-7 PubMed PMID: 38908383.

3. Hoefle-Bénard J, Salloch S. Mass drug administration for neglected tropical disease control and elimination: a systematic review of ethical reasons. BMJ Glob Health. 2024 Mar 13;9(3). doi:10.1136/bmjgh-2023-013439 PubMed PMID: 10.1136/bmjgh-2023-013439.

4. Wolfe CM, Barry A, Campos A, Farham B, Achu D, Juma E, et al. Control, elimination, and eradication efforts for neglected tropical diseases in the World Health Organization African region over the last 30 years: A scoping review. Int J Infect Dis. 2024 Apr 1;141:106943. doi:10.1016/j.ijid.2024.01.010

5. Ca J, P VBK, Kandi V, N G, K S, Dharshini D, et al. Neglected Tropical Diseases: A Comprehensive Review. Cureus. 2024 Feb 9;16. doi:10.7759/cureus.53933

6. Shalla C, Louise NLJ, Njie SS. Bringing the Neglected out of Neglected Tropical Diseases: A Systematic Literature Review. Rehabil Sci. 2025 Sep;10(3):38–42. doi:10.11648/j.rs.20251003.11

7. Lin Y, Long Q, Sun J, Li J, Li Y, Liu J, et al. Global trends, health inequalities, and projections in the burden of neglected tropical diseases and malaria from 1990 to 2021. Sci Rep. 2025 Jul 1;15(1):20958. doi:10.1038/s41598-025-05530-y

8. Ending the neglect to attain the Sustainable Development Goals: A road map for neglected tropical diseases 2021–2030 [Internet]. [cited 2026 May 5]. Available from: https://www.who.int/publications/i/item/9789240010352

9. Towards zero leprosy. Global leprosy (Hansen’s Disease) strategy 2021–2030 [Internet]. [cited 2026 May 5]. Available from: https://www.who.int/publications/i/item/9789290228509

10. Oyeyemi OT, Ogundahunsi O, Schunk M, Fatem RG, Shollenberger LM. Neglected tropical disease (NTD) diagnostics: current development and operations to advance control. Pathog Glob Health. 2024 Jan 2;118(1):1–24. doi:10.1080/20477724.2023.2272095 PubMed PMID: 37872790.

11. Joubert A, Reid M. Knowledge, skills, and training of community health workers to contribute to interprofessional education: a scoping review. J Interprof Care. 2024 Mar 3;38(2):308–18. doi:10.1080/13561820.2023.2176472 PubMed PMID: 36821383.

12. Singh J, Steele K, Singh L. Combining the Best of Online and Face-to-Face Learning: Hybrid and Blended Learning Approach for COVID-19, Post Vaccine, & Post-Pandemic World. J Educ Technol Syst. 2021 Dec 1;50(2):140–71. doi:10.1177/00472395211047865

13. Cano M, Ruiz-Postigo JA, Macharia P, Amoako YA, Phillips RO, Kinyeru E, et al. Evaluating the World Health Organization’s SkinNTDs App as a Training Tool for Skin Neglected Tropical Diseases in Ghana and Kenya: Cross-Sectional Study. J Med Internet Res. 2024 Apr 30;26(1):e51628. doi:10.2196/51628

14. Moungui HC, Iyawa PT, Nana-Djeunga H, Ruiz-Postigo JA, Carrion C. Usability and quality evaluation of the World Health Organization SkinNTDs app among frontline health workers in Cameroon: A mixed methods study. PLoS Negl Trop Dis. 2025 Sep 10;19(9):e0013461. doi:10.1371/journal.pntd.0013461

15. Monitoring the global leprosy situation [Internet]. [cited 2026 May 5]. Available from: https://www.who.int/activities/improving-treatment-for-snakebite-patients

16. Kaba M, Hailemichael Y, Alemu AY, Cherkose T, Kebebew G, Kassa FA, et al. Understanding experiences of neglected tropical diseases of the skin: a mixed-methods study to inform intervention development in Ethiopia. BMJ Glob Health. 2025 Feb 5;10(2). doi:10.1136/bmjgh-2024-016650 PubMed PMID: 10.1136/bmjgh-2024-016650.

17. Chu F. Implementation science: why should we care? J Med Libr Assoc. 2024 Jul 29;112(3):281–5. doi:10.5195/jmla.2024.1919

18. Glasgow RE, Harden SM, Gaglio B, Rabin B, Smith ML, Porter GC, et al. RE-AIM Planning and Evaluation Framework: Adapting to New Science and Practice With a 20-Year Review. Front Public Health. 2019 Mar 29;7. doi:10.3389/fpubh.2019.00064

19. Wilson ME, Ko AI, Reis MG. Collaborative Teaching and Learning: A Model for Building Capacity and Partnerships to Address NTDs. PLoS Negl Trop Dis. 2011 Mar 29;5(3):e939. doi:10.1371/journal.pntd.0000939

20. Rotureau B, Waleckx E, Jamonneau V, Solano P, Molia S, Debré P, et al. Enhancing research integration to improve One Health actions: learning lessons from neglected tropical diseases experiences. BMJ Glob Health. 2022 Jun 10;7(6):e008881. doi:10.1136/bmjgh-2022-008881 PubMed PMID: 35688485; PubMed Central PMCID: PMC9189848.

21. Sharma D, Sood AK, Darius PSH, Gundabattini E, Darius Gnanaraj S, Joseph Jeyapaul A. A Study on the Online-Offline and Blended Learning Methods. J Inst Eng India Ser B. 2022;103(4):1373–82. doi:10.1007/s40031-022-00766-y PubMed PMID: null; PubMed Central PMCID: PMC9252556.

22. Yu Z, Xu W, Sukjairungwattana P. Meta-analyses of differences in blended and traditional learning outcomes and students’ attitudes. Front Psychol. 2022 Sep 16;13. doi:10.3389/fpsyg.2022.926947

23. Paul S, Burman RR, Singh R. Training effectiveness evaluation: Advancing a Kirkpatrick model based composite framework. Eval Program Plann. 2024 Dec 1;107:102494. doi:10.1016/j.evalprogplan.2024.102494

24. Yu JE. Reliability and validity of applying Kirkpatrick model for evaluating exercise rehabilitation program. J Exerc Rehabil. 2025 Aug 31;21(4):200–9. doi:10.12965/jer.2550428.214 PubMed PMID: 40917575; PubMed Central PMCID: PMC12409149.

25. Laronze F, Nkaoua B. Differences between face-to-face and synchronous distance learning on cognitive, behavioural and affective outcomes. Open Learn J Open Distance E-Learn. 2025 Jul 3;40(3):234–51. doi:10.1080/02680513.2024.2399842

26. Fitzpatrick C, Fleming FM, Madin-Warburton M, Schneider T, Meheus F, Asiedu K, et al. Benchmarking the Cost per Person of Mass Treatment for Selected Neglected Tropical Diseases: An Approach Based on Literature Review and Meta-regression with Web-Based Software Application. PLoS Negl Trop Dis. 2016 Dec 5;10(12):e0005037. doi:10.1371/journal.pntd.0005037

27. Godwin-Akpan TG, Diaconu K, Edmiston M, Smith JS, Sosu F, Weiland S, et al. Assessing the cost-effectiveness of integrated case management of Neglected Tropical Diseases in Liberia. BMC Health Serv Res. 2023 Jun 29;23(1):705. doi:10.1186/s12913-023-09685-0

28. Ndikubwimana F, Nirere G, Habimana SS, Akayezu D. Students’ Perceptions on Implementation of Blended Learning Approach at Rwanda Polytechnic-Kigali College. East Afr J Educ Stud. 2026 May 6;9(2):363–78. doi:10.37284/eajes.9.2.4934

29. Zakariya YF, Danlami KB, Shogbesan YO. Affordances and constraints of a blended learning course: experience of pre-service teachers in an African context. Humanit Soc Sci Commun. 2024 Nov 22;11(1):1596. doi:10.1057/s41599-024-04136-5

30. Nemes J. Factors Affecting the Implementation of Blended Learning in Higher Learning Institutions in Tanzania: A Comprehensive Review of Literature. Int J Pedagogy Soc Stud. 2025 Jun 30;10(1):81–98. doi:10.17509/ijposs.v10i1.84732

31. Sareen S, Mandal S. Challenges of blended learning in higher education across global north-south: A systematic and integrative literature review. Soc Sci Humanit Open. 2024 Jan 1;10:101011. doi:10.1016/j.ssaho.2024.101011

